# Validation of motor and functional scales for the evaluation of adult patients with 5q spinal muscular atrophy

**DOI:** 10.1101/2021.06.12.21258357

**Authors:** Juan F Vázquez-Costa, Mónica Povedano, Andrés E Nascimiento-Osorio, Antonio Moreno Escribano, Solange Kapetanovic Garcia, Raul Dominguez, Jessica M Exposito, Laura González, Carla Marco, Julita Medina Castillo, Nuria Muelas, Daniel Natera de Benito, Nancy Carolina Ñungo Garzón, Inmaculada Pitarch Castellano, Teresa Sevilla, David Hervás

**Affiliations:** Neuromuscular Unit, Department of Neurology, Hospital Universitario y Politécnico la Fe, Valencia (Spain); Centro de Investigación Biomédica en Red de Enfermedades Raras (CIBERER), Valencia (Spain); Department of Medicine, Universitat de València, Valencia (Spain); Motor Neuron Unit, Neurology Department, Bellvitge Hospital-IDIBELL; Neuromuscular Unit, Neuropediatric Department, Institut de Recerca Pediàtrica Hospital Sant Joan de Déu, Barcelona, Spain; Center for the Biomedical Research on Rare Diseases (CIBERER), ISCIII, Spain; Neuromuscular Unit, Neurology Department, Hospital Clínico Universitario Virgen de la Arrixaca, Murcia, Spain; ALS and Neuromuscular Unit, Neurology Department, Hospital Universitario Basurto - OSI Bilbao, Spain; Phisycal Medicine and Rehabilitation department. Hospital Sant Joan de Deu. Barcelona; Department of Applied Statistics and Operational Research and Quality, Universitat Politècnica de València, Valencia, Spain

**Keywords:** Spinal Muscular Atrophy, nusinersen, treatment, adults, cohort study

## Abstract

**Objective:** To assess in adult spinal muscular atrophy (SMA) patients the construct validity and responsiveness of several outcome measures.

**Methods:** Patients older than 15 years and followed-up at least for 6 months, between October 2015 and August 2020, with one motor function scale (Hammersmith Functional Motor Scale Expanded, HFMSE; Revised Upper Limb module, RULM) in five referral centers were included. Bedside functional scales (Egen Klassification, EK2; Revised Amyotrophic Lateral Sclerosis Functional Rating Scale, ALSFRS-R) were also collected when available. Correlations and regression models were performed to evaluate the construct validity. The monthly slopes of change were used to calculate their responsiveness.

**Results:** The study included 79 SMA patients, followed up for a mean of 16 months. All scales showed strong or very strong correlations with each other. A floor effect in motor function scales was found in weakest patients (HFMSE < 5 and RULM<10), and a ceiling effect in stronger patients (with HFMSE >55 and RULM > 35), when compared with other scales. ALSFRS-R (B=0.72) showed a strong discriminating ability between walkers, sitters, and non-sitters, and HFMSE (B=0.86) between walkers and sitters. The responsiveness was overall low, although in treated patients a moderate responsiveness was found for ALSFRS-R and HFMSE in walkers (0.69 and 0.61 respectively), and for EK2 in sitters (0.65) and non-sitters (0.60).

**Conclusions:** This study shows the validity in SMA adult patients of commonly used scales. Overall, bedside functional scales showed some advantages over motor function scales, although all scales showed low responsiveness in untreated patients.

## Introduction

5q spinal muscular atrophy (SMA) is a genetic neurodegenerative disease, causing progressive muscular weakness and atrophy, followed by respiratory insufficiency, dysarthria and dysphagia.^1^ According to the age of symptoms onset and to the highest acquired motor milestone, SMA children are typically classified in type 1-3. SMA type 1 patients will never be able to sit unsupported, while SMA type 2 patients will never be able to walk independently.^1^ The rare type 4 patients typically start after 30 years old and will not present any noteworthy disability.^2^ Since SMA is a progressive disease, the SMA type does not reliably inform about the actual functionality in the adulthood. Therefore, adult SMA patients, are preferably functionally classified in non-sitters, sitters and walkers.^1,3^

In the last few years, two genetic-based therapies (nusinersen and risdiplam) have been approved for the treatment of adult SMA patients. However, high quality evidence of their efficacy in this subpopulation is lacking. One major drawback is how to measure changes in adult SMA patients.

On the one hand, motor function scales assess the ability of a patient to perform certain tasks in the clinic, which are used as proxies of day-to-day patients’ functionality.^2^ The Hammersmith Functional Motor Scale Expanded (HFMSE) and the Revised Upper Limb Module (RULM) are probably the most widely used motor scales in late-onset SMA patients.^3–7^ However, they require qualified staff, appropriate facilities and are time-consuming.^2^ Moreover, they have been developed and validated in paediatric populations and its construct validity or sensitivity in adult patients has not been formally assessed.^2,3^ Other motor scales frequently used in adults SMA patients are the muscle strength measurement and the 6 Minute Walk Test (6MWT).^8–13^ Nevertheless, there is no consensus on how to measure the muscle strength and the 6MWT can only be used in ambulant SMA patients.

On the other hand, bedside functional scales measure patients’ disability, based on a rater’s scoring of certain signs or symptoms. Compared with motor function scales, they are usually faster and easier to administer. Consequently, they are frequently used as outcome measures in both clinical practice and research in adult patients with neurodegenerative diseases. The Egen classification 2 (EK2) scale, the revised version of Amyotrophic Lateral Sclerosis Functional Rating Scale (ALSFRS-R) and the SMA Functional Rating Scale (SMAFRS) are the most frequently used bedside functional scales in adult SMA patients.^3^ They are reliable, fast, and easy to use, but data on their construct validity and responsiveness (i.e. sensitivity to change) in adult SMA patients are lacking or scarce.^7,10,12,14^

Therefore, this study aimed to assess the construct validity and responsiveness of a set of motor function (HFMSE, RULM, 6MWT) and bedside functional (EK2 and ALSFRS) scales in adult SMA patients. These properties could help to define their usefulness for both clinical trials and clinical practice.

## Methods

### Study design and participants

For this prospective observational study, SMA patients from 5 centres in Spain were included (Hospital la Fe, Hospital Sant Joan de Deu, Hospital de Bellvitge, Hospital Virgen de la Arrixaca, Hospital de Basurto). The inclusion criteria were: a) genetically confirmed SMA (biallelic mutation in *SMN1*); b) older than 15 years old at the baseline visit; c) data on at least one functional and motor scale at the time of the study closure (August 10^th^ 2020). Some patients were treated with nusinersen according to the routine clinical practice. When available, retrospective data of untreated patients were also collected from October 2015.

### Procedures

Motor and functional scales were administered every 6-12 months by experienced and/or trained neurologists and physiotherapists. Although all centres shared the same protocol, some tests were missing in some visits. Moreover, not all scales are applicable to all patients (see below). Consequently, the number of subjects varies as per scale and visit.

### Clinical variables and outcomes

Age, gender, and age at symptom’s onset and the presence of severe scoliosis (>45º Cobb angle) and/or scoliosis surgery were recorded in all the patients upon recruitment. Patients were classified in type 1 to 4 as defined elsewhere,^2^ as well as in functional subgroups:^1^ walkers (able to walk at least 5 steps without assistance), sitters (able to sit without assistance nor head support for more than 10 seconds) and non-sitters.

For this study, following scales were collected:

1. HFMSE consists of 33 items, with a maximum of 66 points (higher scores indicating better function). It was originally designed for the assessment of high functioning type II and type III SMA patients, that is, sitters and walkers.^15^ It has been validated in SMA children.^16^ Although its content validity and clinical meaningfulness has also been explored in adults,^17^ a significant floor effect has been found in these patients.^13^
2. RULM: It includes 20 items with a maximum score of 37 (higher scores indicating better function).^18^ Although it has been validated in both ambulant and non-ambulant patients, it shows ceiling effect in up to a third of ambulant SMA type 3 patients (without upper limb weakness)^19^ and floor effect at least in a proportion of non-sitters.^6^
3. 6MWT: It measures the distance a patient is able to walk within 6 minutes, and it has been validated in ambulant adult SMA patients.^11^
4. EK2 is a functional scale that includes 17 items on 8 daily-life categories (wheelchair use, wheelchair transfers, trunk mobility, eating, swallowing, breathing, coughing, fatigue). Each item is scored from 0 to 3 for a maximum of 51 points (higher scores indicating worse function). EK2 was designed for non-ambulatory SMA patients, and its convergent validity has been shown in SMA patients with different age ranges, including older adults.^20–22^
5. ALSFRS-R is a functional scale that includes 12 items on 4 domains (bulbar, upper limbs, lower limbs, respiratory). Each item is scored from 0 to 4 for a maximum of 48 points (higher scores indicating better function). It was designed for ALS patients, but it has also been used to assess disability in SMA patients,^10,23^ although a formal validation is lacking.
6. The percent-predicted forced vital capacity (FVC%).

### Statistical analysis

Data were summarised as means, standard deviations, medians, and first and third quartiles for the continuous variables, and as relative and absolute frequencies for the categorical variables. Exploratory descriptive analyses were used to assure the quality of the data.

Convergent validity of the different scales was assessed by means of a correlation matrix, using Spearman’s rho correlations. The strength of correlation was quantified as moderate when rs = 0.50 to 0.69, strong when rs = 0.70 to 0.89 and very strong when rs > 0.90. Scatter plots with trend lines were estimated by local regressions’ analysis to assess possible floor and ceiling effects of the different scales.

For the discriminant validity assessment either logistic (EK2, HFMSE) or ordinal (ALSFRS-R, RULM, FVC%) regressions models were performed for each scale, using as response variable the functional classification (walker, sitter, non-sitter). We then assessed the concordance between the predictions made by these models and the actual classification, using the Bangdiwala’s observer agreement card for ordinal variables.^24^ The agreement was quantified as moderate when B = 0.50 to 0.69, strong when B = 0.70 to 0.89 and very strong when B > 0.90.^24^

The responsiveness of each scale was studied by analyzing their monthly slopes of change between the baseline and last available follow-up, using a linear regression. These slopes were then expressed as standardized response means (SRMs) by calculating the ratios of the mean slopes to their SDs.^25^ Responsiveness was considered low if < 0.50, moderate if ranging from 0.50 to 0.79, and large if > 0.80.^25^

All analyses were pre-specified before looking at the data. P values < 0.05 were considered statistically significant. All the statistical analyses and graphs were performed with the R software (version 4.0.3).

### Ethical approval

The study was approved by the Ethics Committee for Biomedical Research of Instituto de Investigación Sanitaria la Fe and Fundació Sant Joan de Déu. All the participants gave written informed consent.

## Results

### Patients’ characteristics

The study included 79 SMA patients. Their demographic and clinical characteristics are summarised in Table 1. As expected, patients’ functional subgroups differed in their clinical characteristics, although there was a considerable overlap in the SMA types and *SMN2* copy number.

**Table 1.** Demographic and baseline clinical characteristics of SMA patients included in the study. ALSFRS-R: Revised version of the Amyotrophic Lateral Sclerosis Functional Scale; EK2: Egen Klassifikation 2; FVC%: Percent-predicted Forced Vital Capacity; HFMSE: Hammersmith Functional Motor Scale Expanded; RULM: Revised Upper Limb Module; 6MWT: 6-Minutes Walk Test.

| Variable |  |  | Non sitter<br>(n = 30) | Sitter<br>(n = 25) | Walker<br>(n = 24) |
| --- | --- | --- | --- | --- | --- |
| Age (years) |  | Mean (SD) | 26.66 (12.77) | 34.16 (12.71) | 35.84 (14.34) |
|  |  | Median<br>(IQR) | 21.55 (16.83,<br>34.38) | 33.48 (25.14,<br>43.59) | 33.51 (22.23,<br>48.55) |
| Male sex |  | N (%) | 15 (50%) | 8 (32%) | 14 (58.33%) |
| SMA<br>type | 2a | N (%) | 21 (70%) | 1 (4%) | 0 (0%) |
|  | 2b |  | 3 (10%) | 5 (20%) | 0 (0%) |
|  | 3a |  | 4 (13.33%) | 13 (52%) | 6 (25%) |
|  | 3b |  | 2 (6.67%) | 6 (24%) | 15 (62.5%) |
|  | 4 |  | 0 (0%) | 0 (0%) | 3 (12.5%) |
| SMN2<br>copies | 1 | N (%) | 1 (3.33%) | 0 (0%) | 0 (0%) |
|  | 2 |  | 5 (16.67%) | 1 (4%) | 1 (4.17%) |
|  | 3 |  | 22 (73.33%) | 20 (80%) | 8 (33.33%) |
|  | 4 |  | 2 (6.67%) | 4 (16%) | 15 (62.5%) |
| Disease<br>duration<br>(years) |  | Mean (SD) | 25.45 (11.88) | 30.21 (10.64) | 25.19 (16) |
|  |  | Median<br>(IQR) | 20.88 (16.22,<br>33.1) | 27.8 (22.47,<br>39.1) | 23.44 (8.83,<br>37.61) |

Table 1. Demographic and baseline clinical characteristics of SMA patients included in the study.
| NIV use | N (%) |  |  |  |
| --- | --- | --- | --- | --- |
| No |  | 10 (34.48%) | 20 (80%) | 24 (100%) |
| 8h |  | 18 (62.07%) | 5 (20%) | 0 (0%) |
| 24h |  | 1 (3.45%) | 0 (0%) | 0 (0%) |
| Gastrostomy | N (%) | 1 (3.33%) | 0 (0%) | 0 (0%) |
| Severe scoliosis | N (%) | 30 (100%) | 16 (64%) | 1 (4.17%) |
| Nusinersen | N (%) | 10 (33.33%) | 16 (64%) | 13 (54.17%) |
| Salbutamol | N (%) | 20 (66.66%) | 12 (48%) | 9 (37.5%) |
| HFMSE | Mean (SD) | NA (NA) | 10.04 (8.84) | 49.95 (11.54) |
|  | Median (IQR) | NA (NA, NA) | 6.5 (3.75, 16.5) | 53.5 (44.75, 57) |
| RULM | Mean (SD) | 6.69 (6.35) | 19.03 (9.17) | 33.35 (4.95) |
|  | Median (IQR) | 5 (1, 10.12) | 19 (12.5, 25.5) | 36 (30.75, 37) |
| 6MWT | Mean (SD) | NA (N) | NA (NA) | 342.06 (147.27) |
|  | Median (IQR) | NA (NA, NA) | NA (NA, NA) | 362.75 (221.25, 454.5) |
| ALSFRS-R | Mean (SD) | 18 (5.42) | 30.58 (4.14) | 42.25 (2.87) |
|  | Median (IQR) | 19 (16, 21) | 31 (28, 32.5) | 42.5 (40, 43.75) |
| EK2 | Mean (SD) | 26.88 (8.75) | 14.31 (6.92) | NA (NA) |
|  | Median<br>(IQR) | 26.5 (21.25,<br>32.5) | 14 (8.75, 19.25) | NA (NA, NA) |
| FVC% | Mean (SD) | 32.42 (17.06) | 76.55 (37.62) | 101.11 (17.23) |
|  | Median<br>(IQR) | 31.6 (18.5, 40.5) | 70.2 (49, 105) | 102.5 (88.78,<br>110.75) |
| Follow-up<br>(months) | Mean (SD) | 16.19 (9.66) | 14.95 (5.45) | 16.64 (7.67) |
|  | Median<br>(IQR) | 15.17 (9.8, 22.1) | 14.47 (11.43,<br>17.73) | 15.3 (12.07,<br>21.68) |

### Convergent validity

All motor function and bedside functional scales showed either strong or very strong correlation with each other (Figure 1). The greatest correlations were between EK2 and ALSFRS-R (rs = −0.96) and between HFMSE and ALSFRS-R (rs = 0.89). The weakest correlations were between FVC and HFMSE (rs = 0.5) and between FVC and 6MWT (rs = −0.04).

**Figure 1.**
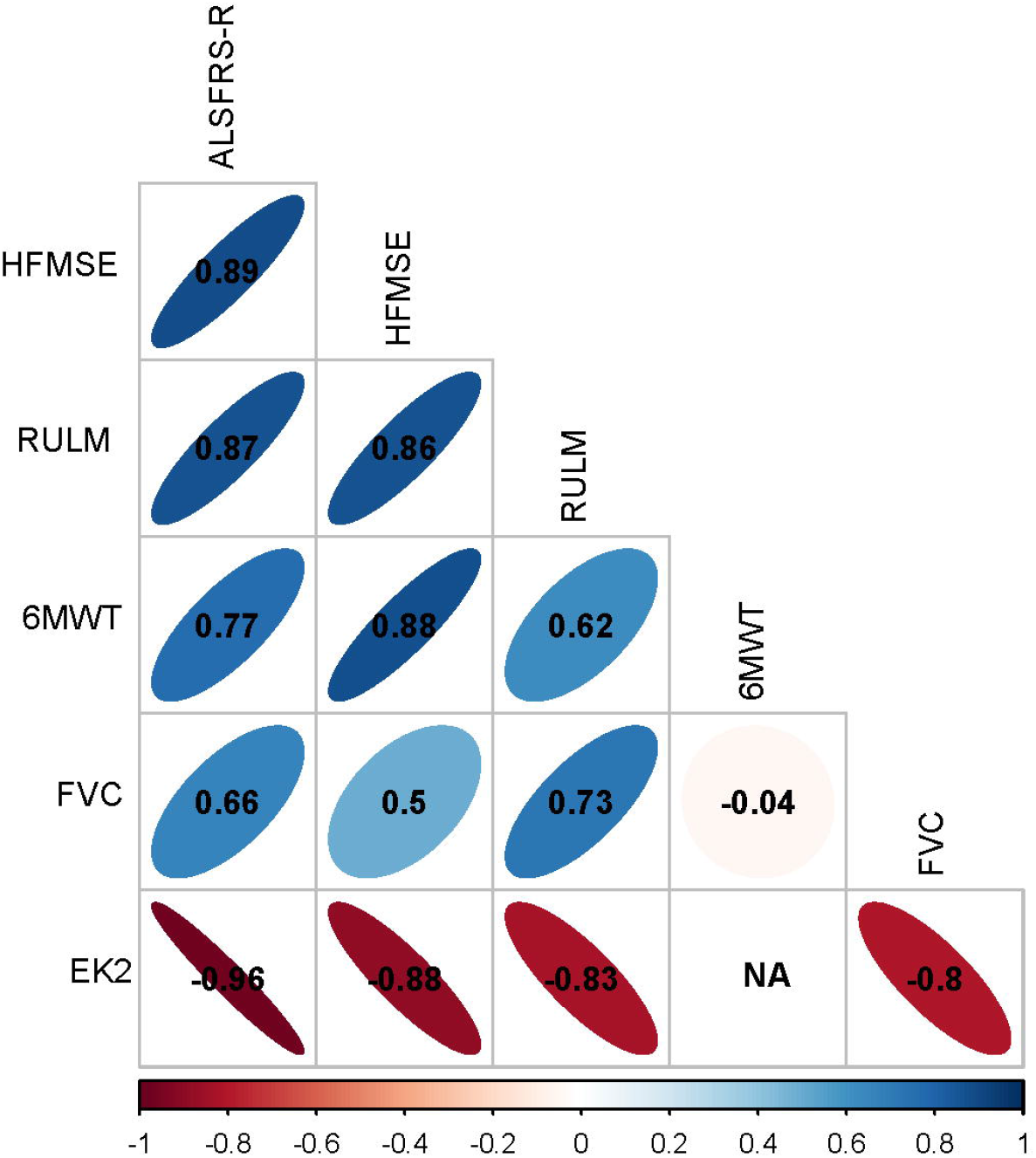
Graphical representation of correlations between outcome measures. Colors represent the strength of correlations and numbers correspond to Spearman’s rho correlations.

Compared with ALSFRS-R, EK2 and RULM, a floor effect of HFMSE was found in weakest sitters (HFMSE < 5, Figure 2A and 2B and 2G). Moreover, a ceiling effect was apparent for walkers with HFMSE >55, when compared with 6MWT (Figure 2C). Regarding RULM, a floor effect was found in patients with RULM < 10, when compared with ALSFRS-R and EK2 (Figure 2D and 2E); and a ceiling effect in patients with RULM > 35 when compared with ALSFRS-R, 6MWT and HFMSE (Figure 2D, 2F and 2G). ALSFRS-R and EK2 showed no apparent floor or ceiling effect, when compared with each other (Figure 2H) or with motor scales (Figure 2A, 2B, 2D, 2E and 2I).

**Figure 2.**
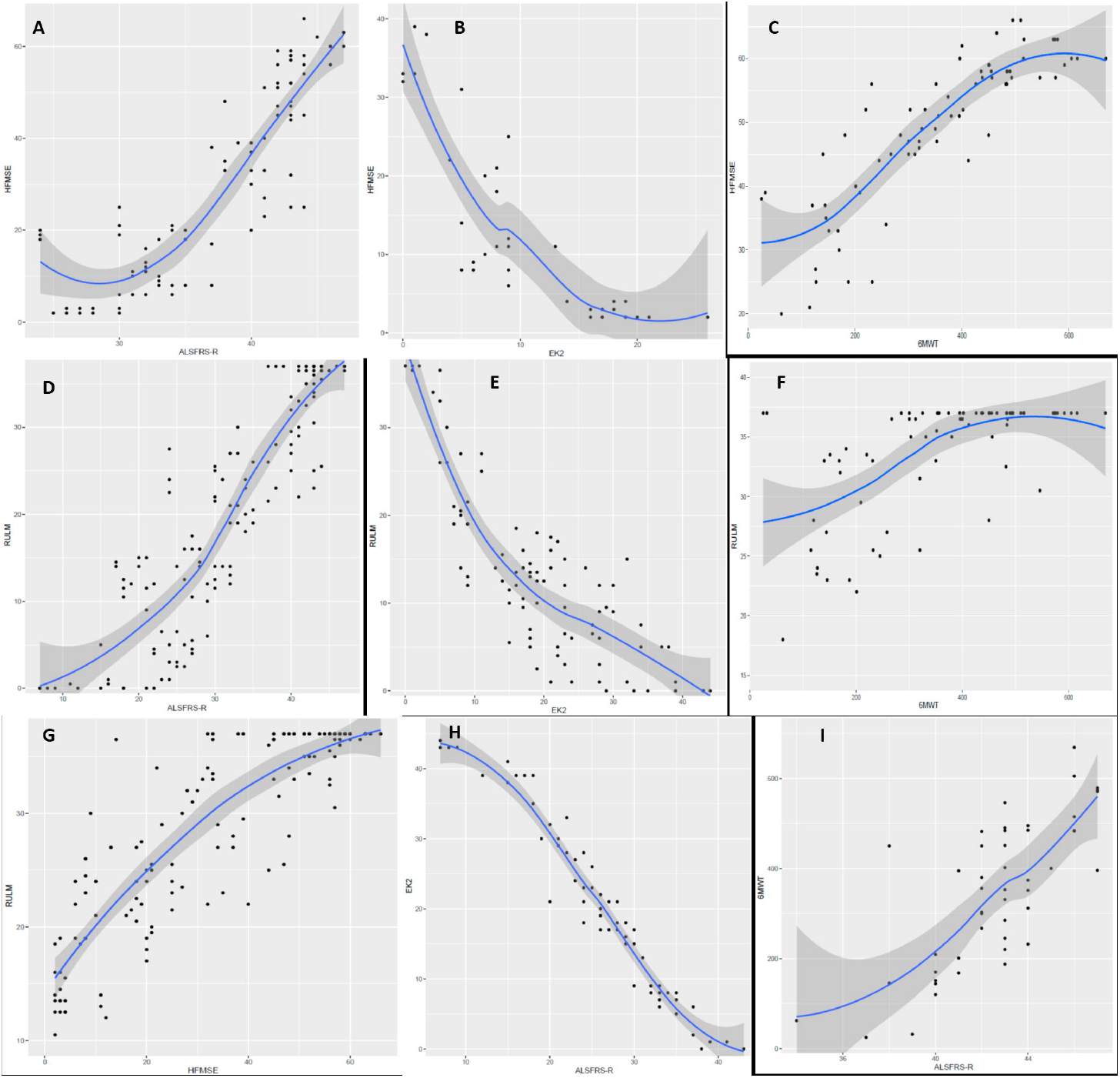
Correlations of HFMSE and ALSFRS-R (A); HFMSE and EK2 (B); HFMSE and 6MWT (C); RULM and ALSFRS-R (D); RULM and EK2 (E); RULM and 6MWT (F); RULM and HFMSE (G); EK2 and ALSFRS-R (H); 6MWT and ALSFRS-R (I). Scatter plots with trend lines were estimated by local regression to analyse possible floor and ceiling effects of the different scales. ALSFRS-R: Revised version of the Amyotrophic Lateral Sclerosis Functional Scale; EK2: Egen Klassifikation 2; HFMSE: Hammersmith Functional Motor Scale Expanded; RULM: Revised Upper Limb Module; 6MWT: 6-Minutes Walk Test.

### Discriminant validity

All scales discriminated between functional subgroups, although with considerable overlap in most of them. Among those tests applicable to all subgroups of patients, only ALSFRS-R showed a strong discriminating ability (B = 0.72, Figure 3A). RULM showed moderate discriminating ability (B = 0.62, Figure 3B) and FVC low (B = 0.35, Figure 3C). Among those scales applicable only to two subgroups of patients, HFMSE showed strong discriminating ability between walkers and sitters (B = 0.86, Figure 3D) and EK2 moderate between sitters and non-sitters (B = 0.68, Figure 3E).

**Figure 3.**
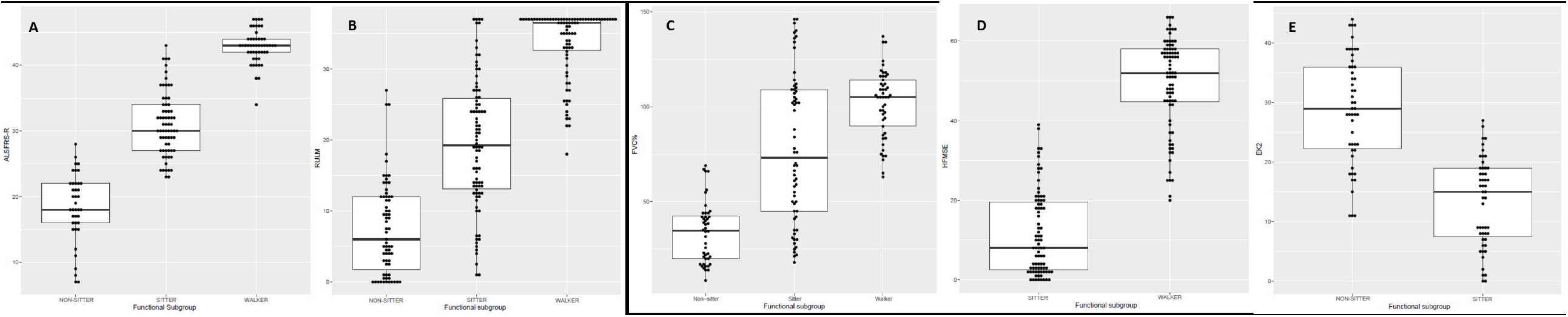
Boxplots of ALSFRS-R (A), RULM (B), FVC% (C), HFMSE (D), and EK2 (E), according to functional subgroups to represent the discriminating ability. ALSFRS-R: Revised version of the Amyotrophic Lateral Sclerosis Functional Scale; EK2: Egen Klassifikation 2; FVC%: Percent-predicted Forced Vital Capacity; HFMSE: Hammersmith Functional Motor Scale Expanded; RULM: Revised Upper Limb Module.

### Responsiveness

Both treated and untreated patients were followed up for a mean of 16 months. In untreated patients the responsiveness was overall low (Table 2): in walkers no measure appeared to adequately capture worsening during follow up, while ALSFRS-R was the most responsive measure in sitters (−0.43) and FVC in non-sitters (−0.37). In treated patients, the responsiveness was overall low too, with some exceptions (Table 3). Moderate responsiveness was found for ALSFRS-R and HFMSE in walkers (0.69 and 0.61 respectively), and for EK2 in sitters (0.65) and non-sitters (0.60).

**Table 2.** Standardized response means of each scale in untreated patients, globally and by functional subgroup. ALSFRS-R: Revised version of the Amyotrophic Lateral Sclerosis Functional Scale; EK2: Egen Klassifikation 2; FVC%: Percent-predicted Forced Vital Capacity; HFMSE: Hammersmith Functional Motor Scale Expanded; RULM: Revised Upper Limb Module; 6MWT: 6-Minutes Walk Test.

| Untreated patients | RULM<br>(n = 36) | HFMSE<br>(n = 21) | 6MWT<br>(n = 10) | EK2<br>(n = 24) | ALSFRS-R<br>(n = 19) | FVC%<br>(n = 33) |
| --- | --- | --- | --- | --- | --- | --- |
| Global | -0.13 | 0.15 | 0.25 | 0.24 | -0.19 | -0.39 |
| Walker | -0.03 | 0.27 | 0.25 | NA | 0.37 | NA |
| Sitter | -0.13 | 0.04 | NA | 0.21 | -0.43 | -0.16 |
| Non-sitter | -0.18 | NA | NA | 0.25 | -0.30 | -0.37 |

**Table 3.**
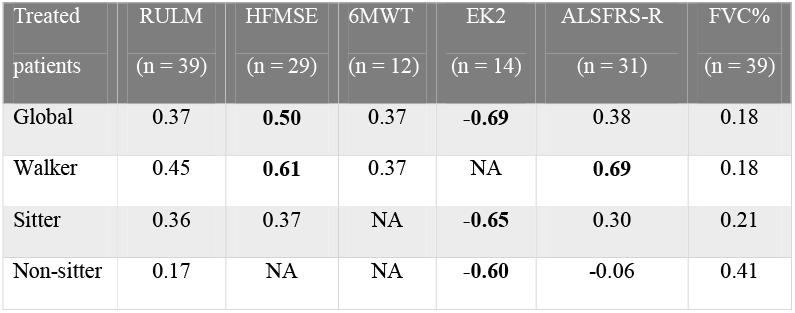
Standardized response means of each scale in treated patients, globally and by functional subgroup. In bold, those scales showing moderate responsiveness. ALSFRS-R: Revised version of the Amyotrophic Lateral Sclerosis Functional Scale; EK2: Egen Klassifikation 2; FVC%: Percent-predicted Forced Vital Capacity; HFMSE: Hammersmith Functional Motor Scale Expanded; RULM: Revised Upper Limb Module; 6MWT: 6-Minutes Walk Test.

## Discussion

In this longitudinal study, we addressed for the first time the validation of a set of motor and functional scales frequently used in the clinical practice to evaluate both the natural history and the efficacy of nusinersen in adult SMA patients.^5,6,10,26^ Until recently, adult SMA patients had been frequently disengaged and neglected from both health services and research studies.^27–29^ The approval of disease-modifying treatments has resulted in a substantial rise of referrals, and an increasing interest of research in adult SMA patients.^29,30^ At present, motor function scales most frequently used in adult SMA patients have been designed and validated in children. However, both populations present considerable differences (e.g., in disease progression rate, contractures and scoliosis, comorbidities, etc.…) that could diversely affect the performance of motor scales. Moreover, few data on functional scales are available in SMA, despite they are frequent outcome measures in both clinical practice and research in most neurodegenerative diseases.^31^

Both valid and responsive scales are warranted to improve research in adult SMA patients. Therefore, we used longitudinal data of a multicentre cohort of adult SMA patients to assess the main properties of motor function and bedside functional scales, which will be discussed hereafter.

### HFMSE

HFMSE was designed to evaluate type 2 and 3 SMA patients and has been widely used in clinical practice and research. Although its content validity and clinical meaningfulness has been shown in adult SMA patients,^17^ its construct validity has not been formally assessed in adult SMA patients. Consistently with previous results in children,^14,16,32^ we found that HFMSE strongly correlates with other motor function and bedside functional scales, and discriminated well between sitters and walkers, suggesting its convergent and discriminant validity also in adult patients. However, it correlated only moderately with FVC%, suggesting less convergent validity in more severely affected patients (weakest sitters). Indeed, it showed a floor effect in weakest sitters (HFMSE < 5) when compared with other motor and functional scales (ALSFRS-R, EK2 and RULM). Interestingly, a similar floor effect has been recently described in type 2 and type 3a adults, when compared with the muscle strength.^13^ Moreover, we found some ceiling effect in highly functioning walkers (HFMSE > 60), when HFMSE was compared with 6MWT. Remarkably, the floor and ceiling effect together with some pitfalls in their psychometric properties (such as a strong differential item functioning with age) had been previously suggested and led to a new version, the Revised Hammersmith Scale (RHS).^33,34^ Finally, we found a moderate responsiveness in treated walkers, suggesting that it could be useful to detect the effect of new treatments in this subgroup of patients, as has been previously shown.^5,6^

### RULM

RULM was designed with the help of Rasch analysis, to evaluate upper limb function in type 2 and 3 SMA patients.^18^ Its convergent validity was demonstrated by showing strong correlation with HFMSE, ALSFRS-R, EK2 and FVC in a study with type 2 and 3 SMA patients of mixed ages, but mostly children.^19^ However, it correlated only moderately with 6MWT, suggesting less convergent validity in strong walkers. We confirmed these findings in adult patients and found a ceiling effect in patients with RULM > 35, when compared with ALSFRS-R, HFMSE and 6MWT. Moreover, a mild floor effect was also found in weakest patients scoring <10 in RULM, when compared with ALSFRS-R and EK2. RULM showed only moderate discriminant validity, since sitters scores were very variable. Its responsiveness after 16 months of follow up was overall low in all patients, confirming that few changes can be expected in short time periods (less than two years),^19^ even after treatment with nusinersen.

### 6MWT

This study confirms the convergent validity of 6MWT in ambulant adult SMA patients, which had been recently suggested.^11^ Moreover, it has been found useful to detect improvements in motor function and fatigability after nusinersen treatment.^5,6,10,35^ However, the 6MWT is only applicable to walkers. Furthermore, its responsiveness and reliability have been found to be lower than other clinical outcome measures in this and previous studies,^12,26,36^ probably limiting its utility, when compared with other measures.

### EK2

The construct validity of EK2 has been previously suggested in a study including a large proportion of adults.^37^ This study confirms its convergent validity, by showing strong correlations with HFMSE, RULM, ALSFRS-R and FVC%. Moreover, neither floor nor ceiling effect was found when compared with these scales. It also showed a moderate discriminating ability between sitters and non-sitters. Remarkably, it showed a consistent responsiveness in both sitters and non-sitters, which was moderate in the case of treated patients. This is in agreement with previous studies, which showed EK2 ability to detect treatment-related changes in non-ambulant patients.^14^ Despite these encouraging data, a Rasch analysis found some pitfalls in this scale that must be considered.^34^

### ALSFRS-R

Although not formally validated, ALSFRS-R has been widely used to assess disability in adult SMA patients,^10,23,38^ suggesting its content validity in them. Here, we demonstrate its construct validity. Strong correlations were found with all motor and functional scales, and neither floor nor ceiling effect was apparent, when compared with them. Furthermore, it showed a strong discriminating ability and a moderate responsiveness in treated walkers. This is in contrast with previous studies,^10,38^ which failed to detect improvements in ALSFRS-R in ambulant adult SMA patients after nusinersen treatment. However, these studies were probably underpowered since they included only a few patients with short follow-up.

### FVC%

The convergent validity of FVC% has been previously assessed in a large cohort of SMA patients, mostly children.^39^ This study confirms moderate correlations of FVC% with functional scales, albeit its discriminating ability and responsiveness was overall low. However, it may still be useful in non-sitters, in whom its responsiveness is higher and respiratory endpoints are particularly clinically meaningful.

### Selecting the best outcome measures

Selecting the best outcome measures in adult SMA patients is of the utmost importance to assess the efficacy of new treatments in both the clinical practice and clinical trials. However, the huge heterogeneity of adult patients (in both age and function), the slow decline rate and the scarcity of natural history studies are major challenges to be considered. All scales analysed here have their strengths and limitations. However, bedside functional scales have some advantages over motor function scales. Firstly, in comparison to most motor function scales, they are faster, cheaper and easier to administrate. Secondly, they can encompass and distinguish a great range of functional states, reducing the floor and ceiling effects of motor scales, as shown above. Thirdly, important items that are not covered in motor scales (e.g., fatigue and bulbar or respiratory problems) are addressed in functional scales. Fourthly, self and telematic administration of functional scales such as ALSFRS-R have been shown reliable and reproducible,^40^ facilitating patients’ follow up. Finally, bedside functional scales provide a unique insight into the clinical relevance of a score change at an individual level, which could be only inferred with motor scales.

Accordingly, an overall good validity and responsiveness has been found in bedside functional scales, when compared with motor outcomes in this and other pilot studies in adult SMA patients.^11,12,37,41^ However, they are not without limitations. Reliability problems might appear if items are not clearly delimited. Moreover, some pitfalls, such as multidimensionality, have been found after a Rasch analysis of EK2 and ALSFRS-R.^34,42^ Nevertheless, these limitations can be addressed with the use of different statistical approaches,^43^ or the development of new functional scales.^42^

Regarding motor function scales, those designed and validated in SMA children (such as HFMSE and RULM) showed overall acceptable construct validity also in adult patients. However, when compared with functional scales, they showed floor and ceiling effects in patients at the ends of the clinical spectrum. They also showed lower responsiveness in this and another study.^12^ Moreover, most of them are time-consuming, require training and infrastructure, and are probably not useful in all functional subgroups. Overall, both motor and functional scales showed low sensitivity in untreated patients to detect changes after a median of 16 months. This is not surprising since SMA is a very slowly progressing disease in the adulthood. Interestingly, quantitative and semi-quantitative strength measures have shown very promising results in ambulant and non-ambulant adult SMA patients in natural history studies.^11–13^ These and the recently developed RHS^33^ warrant further studies in the adult population.

Future studies should also assess the minimal detectable change and minimal clinically important change of the different scales.^44^ These parameters can help to quantify individual responses to treatment, guiding decisions about treatment discontinuation in the clinical practice or serving as endpoints in clinical trials.

Given the complexity of measuring changes in adult SMA patients, we believe that the combined use of several outcome measures will be needed. Ideally, all measures should be applicable to all functional subgroups of patients (for example pinch strength, ALSFRS-R or FVC), unless the research is focused in one or two subgroups. Moreover, since each test may be more responsive in a different functional subgroup, stratification is recommended whenever possible. Finally, both patient reported outcomes^45^ and biomarkers should probably be incorporated into the research protocols. Thus, the use of composite multimodal scores is an interesting approach that has been already tested.^12^

### Strengths and limitations

The main strength of this study is the thorough evaluation of adult SMA patients that allowed us to validate several scales simultaneously. However, it also has several limitations, which are common in real-world studies in rare diseases. A greater sample size would have been desirable for the stratified results of responsiveness, especially for bedside functional scales and FVC% that were not routinely collected in all centers. Moreover, not all patients were visited at the same intervals and baseline patients’ characteristics were somewhat different in treated and untreated groups. All this can affect the responsiveness and, consequently, these results must be interpreted with caution. However, the statistical analysis was designed to minimize these limitations, for example by calculating the responsiveness based on the slopes of change.

In conclusion, in this multicenter study we showed the validity in SMA adults of those scales most used to assess late-onset SMA. Overall, bedside functional scales showed some advantages over motor scales, although all analyzed scales showed limited responsiveness. New outcome measures should probably be developed and/or validated in adult SMA patients. Meanwhile, this study provides a framework for the selection of the most relevant scales in the evaluation of adult SMA patients in both the clinical practice and research.

## Data Availability

JFVC and DH had full access to the database population used to create the study population. All data supporting our findings are available on reasonable request.

## Study funding

This study has received funding from FUNDAME (FUN-000-2017-01), from CUIDAME (PIC188-18), from Instituto de Salud Carlos III (JR19/00030 PI JFVC, 19/01178 PI TS), and from Generalitat Valenciana (PROMETEO/ 2018/135, PI TS). The Centro de Investigación Biomédica en Red de Enfermedades Raras (CIBERER) is initiative from the ISCIII. TS and JFVC are members of the European Reference Network for Rare Neuromuscular Diseases (ERN EURO-NMD). Sponsors did not participate in the study design, data acquisition and analysis, data interpretation or in writing the article.

## Disclosures

This study has received funding from FUNDAME (FUN-000-2017-01) and CUIDAME (PIC188-18).

Dr. Vázquez-Costa is funded by grants of the Instituto de Salud Carlos III (JR19/00030, PI Vázquez), and received personal fees from Biogen and Roche outside the submitted work.

Dr.Nascimento-Osorio received personal fees from Avexis, Biogen and Roche outside the submitted work; principal investigator for ongoing Biogen and Roche clinical trials.

Dr. N. Muelas received personal fees from Biogen outside the submitted work.

Dr. A. Moreno received personal fees from Biogen outside the submitted work.

Dr. M Povedano received personal fees from Biogen and Roche outside the submitted work.

Dr Solange Kapetanovic Garcia has nothing to disclose.

Dr Raul Dominguez has nothing to disclose.

Dr Jessica M Exposito has nothing to disclose.

Dr Laura González has nothing to disclose.

Dr Carla Marco has nothing to disclose.

Dr Julita Medina Castillo has nothing to disclose.

Dr Daniel Natera de Benito has nothing to disclose.

Dr Nancy Carolina Ñungo Garzón has nothing to disclose.

Dr. Pitarch-Castellano received personal fees from Avexis, Biogen and Roche outside the submitted work; principal investigator for ongoing Biogen clinical trial.

Dr David Hervás has nothing to disclose.

## Data availability

| **Name** | **Location** | **Contribution** |
| --- | --- | --- |
| Juan F Vázquez-Costa | Hospital Universitario y Politécnico la Fe, Valencia | Designed the study, participated in clinical data acquisition and interpretation, and wrote and edited the manuscript |
| Mónica Povedano | Bellvitge Hospital-IDIBELL | Clinical data acquisition and interpretation, and critically revised the |
|  |  | manuscript |
| Andrés<br>Nascimento-Osorio | Hospital Sant Joan<br>de Déu, Barcelona | Clinical data acquisition and<br>interpretation, and critically revised the<br>manuscript |
| Antonio Moreno<br>Escribano | Hospital Clínico<br>Universitario Virgen<br>de la Arrixaca,<br>Murcia, Spain | Clinical data acquisition and<br>interpretation, and critically revised the<br>manuscript |
| Solange<br>Kapetanovic Garcia | Hospital Universitario<br>Basurto - OSI Bilbao,<br>Spain | Clinical data acquisition and<br>interpretation, and critically revised the<br>manuscript |
| Raul Dominguez | Bellvitge Hospital-IDIBELL | Clinical data acquisition |
| Jessica M Exposito | Hospital Sant Joan de<br>Déu, Barcelona | Clinical data acquisition and<br>interpretation, and critically revised the<br>manuscript |
| Laura González | Bellvitge Hospital- | Clinical data acquisition |
|  | IDIBELL |  |
| Carla Marco | Bellvitge Hospital-IDIBELL | Clinical data acquisition |
| Julita Medina<br>Castillo | Hospital Sant Joan de<br>Déu, Barcelona | Clinical data acquisition and<br>interpretation, and critically revised the<br>manuscript |
| Nuria Muelas | Hospital Universitario<br>y Politécnico la Fe,<br>Valencia | Critically revised the manuscript |
| Daniel Natera | Hospital Sant Joan<br>de Déu, Barcelona | Clinical data acquisition and<br>interpretation, and critically revised the<br>manuscript |
| Nancy Carolina<br>Ñungo Garzón | Hospital Universitario<br>y Politécnico la Fe,<br>Valencia | Clinical data acquisition and<br>interpretation, and critically revised the<br>manuscript |
| Inmaculada<br>Pitarch-Castellano | Hospital Universitario<br>y Politécnico la Fe, | Clinical data acquisition and<br>interpretation, and critically revised the |
|  | Valencia | manuscript |
| Teresa Sevilla | Hospital Universitario<br>y Politécnico la Fe,<br>Valencia | Clinical data acquisition and<br>interpretation, and critically revised the<br>manuscript |
| David Hervás | Universitat<br>Politècnica de<br>València, Valencia | Planned and performed statistical analysis<br>and interpretation, and critically revised<br>the manuscript |

## Notes

### Competing Interest Statement

This study has received funding from FUNDAME (FUN-000-2017-01) and CUIDAME (PIC188-18).
Dr. Vazquez-Costa is funded by grants of the Instituto de Salud Carlos III (JR19/00030, PI Vazquez), and received personal fees from Biogen and Roche outside the submitted work.
Dr.Nascimento-Osorio received personal fees from Avexis, Biogen and Roche outside the submitted work; principal investigator for ongoing Biogen and Roche clinical trials.
Dr. N. Muelas received personal fees from Biogen outside the submitted work.
Dr. A. Moreno received personal fees from Biogen outside the submitted work.
Dr. M Povedano received personal fees from Biogen and Roche outside the submitted work.
Dr Solange Kapetanovic Garcia has nothing to disclose.
Dr Raul Dominguez has nothing to disclose.
Dr Jessica M Exposito has nothing to disclose.
Dr Laura Gonzalez has nothing to disclose.
Dr Carla Marco has nothing to disclose.
Dr Julita Medina Castillo has nothing to disclose.
Dr Daniel Natera de Benito has nothing to disclose.
Dr Nancy Carolina Nungo Garzon has nothing to disclose.
Dr. Pitarch-Castellano received personal fees from Avexis, Biogen and Roche outside the submitted work; principal investigator for ongoing Biogen clinical trial.
Dr David Hervas has nothing to disclose.

### Author Declarations

The study was approved by the Ethics Committee for Biomedical Research of Instituto de Investigacion Sanitaria la Fe and Fundacio Sant Joan de Deu. All the participants gave written informed consent.

## References

1. Mercuri E, Finkel RS, Muntoni F, et al. Diagnosis and management of spinal muscular atrophy: Part 1: Recommendations for diagnosis, rehabilitation, orthopedic and nutritional care. Neuromuscul Disord. 2018;28:103–115.

2. Finkel R, Bertini E, Muntoni F, Mercuri E. 209th ENMC International Workshop: Outcome Measures and Clinical Trial Readiness in Spinal Muscular Atrophy 7-9 November 2014, Heemskerk, The Netherlands. Neuromuscul Disord [online serial]. Elsevier Ltd; 2015;25:593–602. Accessed at: https://pubmed.ncbi.nlm.nih.gov/26045156/. Accessed January 9, 2021.

3. Sansone VA, Walter MC, Attarian S, et al. Measuring Outcomes in Adults with Spinal Muscular Atrophy - Challenges and Future Directions - Meeting Report. J Neuromuscul Dis. 2020;7:523–534.

4. Finkel R, Bertini E, Muntoni F, Mercuri E. 209th ENMC International Workshop: Outcome Measures and Clinical Trial Readiness in Spinal Muscular Atrophy 7-9 November 2014, Heemskerk, The Netherlands. Neuromuscul Disord. Elsevier Ltd; 2015;25:593–602.

5. Hagenacker T, Wurster CD, Günther R, et al. Nusinersen in adults with 5q spinal muscular atrophy: a non-interventional, multicentre, observational cohort study. Lancet Neurol. Lancet Publishing Group; 2020;19:317–325.

6. Maggi L, Bello L, Bonanno S, et al. Nusinersen safety and effects on motor function in adult spinal muscular atrophy type 2 and 3. J Neurol Neurosurg Psychiatry [online serial]. BMJ Publishing Group; 2020;91:1166–1174. Accessed at: http://jnnp.bmj.com/. Accessed November 16, 2020.

7. Yeo CJJ, Simeone SD, Townsend EL, Zhang RZ, Swoboda KJ. Prospective Cohort Study of Nusinersen Treatment in Adults with Spinal Muscular Atrophy. J Neuromuscul Dis. 2020;7:257–268.

8. De Wel B, Goosens V, Sobota A, et al. Nusinersen treatment significantly improves hand grip strength, hand motor function and MRC sum scores in adult patients with spinal muscular atrophy types 3 and 4. J Neurol [online serial]. Springer Science and Business Media Deutschland GmbH; Epub 2020. Accessed at: https://pubmed.ncbi.nlm.nih.gov/32935160/. Accessed November 16, 2020.

9. Miller RG, Moore DH, Dronsky V, et al. A placebo-controlled trial of gabapentin in spinal muscular atrophy. J Neurol Sci [online serial]. J Neurol Sci; 2001;191:127–131. Accessed at: https://pubmed.ncbi.nlm.nih.gov/11677003/. Accessed January 9, 2021.

10. Walter MC, Wenninger S, Thiele S, et al. Safety and Treatment Effects of Nusinersen in Longstanding Adult 5q-SMA Type 3 - A Prospective Observational Study. J Neuromuscul Dis [online serial]. Epub 2019 Sep 28.:1– 13. Accessed at: https://www.medra.org/servlet/aliasResolver?alias=iospress&doi=10.3233/JND-190416. Accessed October 12, 2019.

11. Elsheikh B, King W, Peng J, et al. Outcome measures in a cohort of ambulatory adults with spinal muscular atrophy. Muscle and Nerve. 2020;61:187–191.

12. Querin G, Lenglet T, Debs R, et al. Development of new outcome measures for adult SMA type III and IV: a multimodal longitudinal study. J Neurol [online serial]. Springer Science and Business Media Deutschland GmbH; Epub 2021. Accessed at: https://pubmed.ncbi.nlm.nih.gov/33388927/. Accessed January 9, 2021.

13. Wijngaarde CA, Stam M, Otto LAM, et al. Muscle strength and motor function in adolescents and adults with spinal muscular atrophy. Neurology [online serial]. NLM (Medline); 2020;95:e1988–e1998. Accessed at: https://n.neurology.org/content/95/14/e1988. Accessed February 18, 2021.

14. Frongia AL, Natera-De Benito D, Ortez C, et al. Salbutamol tolerability and efficacy in patients with spinal muscular atrophy type II. Neuromuscul Disord [online serial]. 2019;29:517–524. Accessed at: http://www.sciencedirect.comwww.elsevier.com/locate/nmd. Accessed May 9, 2020.

15. O’Hagen JM, Glanzman AM, McDermott MP, et al. An expanded version of the Hammersmith Functional Motor Scale for SMA II and III patients. Neuromuscul Disord [online serial]. Neuromuscul Disord; 2007;17:693–697. Accessed at: https://pubmed.ncbi.nlm.nih.gov/17658255/. Accessed January 6, 2021.

16. Glanzman AM, O’Hagen JM, McDermott MP, et al. Validation of the Expanded Hammersmith Functional Motor Scale in spinal muscular atrophy type II and III. J Child Neurol [online]. J Child Neurol; 2011. p. 1499–1507. Accessed at: https://pubmed.ncbi.nlm.nih.gov/21940700/. Accessed January 6, 2021.

17. Pera MC, Coratti G, Forcina N, et al. Content validity and clinical meaningfulness of the HFMSE in spinal muscular atrophy. BMC Neurol [online serial]. BMC Neurology; 2017;17:1–10. Accessed at: http://dx.doi.org/10.1186/s12883-017-0790-9.

18. Mazzone ES, Mayhew A, Montes J, et al. Revised upper limb module for spinal muscular atrophy: Development of a new module. Muscle and Nerve [online serial]. John Wiley and Sons Inc.; 2017;55:869–874. Accessed at: https://onlinelibrary.wiley.com/doi/abs/10.1002/mus.25430. Accessed September 16, 2020.

19. Pera MC, Coratti G, Mazzone ES, et al. Revised upper limb module for spinal muscular atrophy: 12 month changes. Muscle and Nerve. 2019;59:426–430.

20. Fagoaga J, Girabent-Farrés M, Bagur-Calafat C, Febrer A, Steffensen BF. Evaluación funcional para personas no ambulantes afectas de atrofia muscular espinal y distrofia muscular de Duchenne. Traducción y validación de la escala Egen Klassifikation 2 para la población espaÑola. Rev Neurol. 2015;60:439–446.

21. Werlauff U, Fynbo Steffensen B. The applicability of four clinical methods to evaluate arm and hand function in all stages of spinal muscular atrophy type II. Disabil Rehabil [online serial]. 2014;36:2120–2126. Accessed at: http://www.tandfonline.com/doi/full/10.3109/09638288.2014.892157. Accessed August 20, 2019.

22. Steffensen BF, Mayhew; A, Aloysius; A, et al. Egen classification revisited in SMA. Neromuscul Disord. 2008;18:84.

23. Wurster CD, Steinacker P, Günther R, et al. Neurofilament light chain in serum of adolescent and adult SMA patients under treatment with nusinersen. J Neurol [online serial]. Springer Berlin Heidelberg; 2020;267:36–44. Accessed at: https://doi.org/10.1007/s00415-019-09547-y.

24. MuÑoz SR, Bangdiwala SI. Interpretation of Kappa and B statistics measures of agreement. J Appl Stat. 1997;24:105–112.

25. Husted JA, Cook RJ, Farewell VT, Gladman DD. Methods for assessing responsiveness: A critical review and recommendations. J Clin Epidemiol [online serial]. Elsevier Inc.; 2000;53:459–468. Accessed at: https://pubmed.ncbi.nlm.nih.gov/10812317/. Accessed January 9, 2021.

26. Yeo CJJ, Simeone SD, Townsend EL, Zhang RZ, Swoboda KJ. Prospective Cohort Study of Nusinersen Treatment in Adults with Spinal Muscular Atrophy. J Neuromuscul Dis. IOS Press; 2020;7:257–268.

27. Wan HWY, Carey KA, D’Silva A, et al. Health, wellbeing and lived experiences of adults with SMA: A scoping systematic review [online]. Orphanet J. Rare Dis. BioMed Central Ltd.; 2020. Accessed at: https://pubmed.ncbi.nlm.nih.gov/32164772/. Accessed October 8, 2020.

28. Wan HWY, Carey KA, D’Silva A, Kasparian NA, Farrar MA. “Getting ready for the adult world”: How adults with spinal muscular atrophy perceive and experience healthcare, transition and well-being. Orphanet J Rare Dis [online serial]. BioMed Central Ltd.; 2019;14. Accessed at:/pmc/articles/PMC6446316/?report=abstract. Accessed November 5, 2020.

29. Walter MC, Chiriboga C, Duong T, et al. Improving Care and Empowering Adults Living with SMA: A Call to Action in the New Treatment Era. J Neuromuscul Dis [online serial]. J Neuromuscul Dis; Epub 2021 Feb 24.:1–21. Accessed at: https://www.medra.org/servlet/aliasResolver?alias=iospress&doi=10.3233/JND-200611. Accessed March 14, 2021.

30. Sansone VA, Coratti G, Pera MC, et al. Sometimes they come back: New and old spinal muscular atrophy adults in the era of nusinersen. Eur J Neurol [online serial]. Blackwell Publishing Ltd; Epub 2020. Accessed at: https://pubmed.ncbi.nlm.nih.gov/33012052/. Accessed January 5, 2021.

31. Vázquez-Costa JF. Natural history data in adults with SMA. Lancet Neurol [online serial]. Elsevier Ltd; 2020;19:564–565. Accessed at: http://dx.doi.org/10.1016/S1474-4422(20)30183-6.

32. O’Hagen JM, Glanzman AM, McDermott MP, et al. An expanded version of the Hammersmith Functional Motor Scale for SMA II and III patients. Neuromuscul Disord [online serial]. Elsevier; 2007;17:693–697. Accessed at: http://www.nmd-journal.com/article/S0960896607001861/fulltext. Accessed September 16, 2020.

33. Ramsey D, Scoto M, Mayhew A, et al. Revised Hammersmith Scale for spinal muscular atrophy: A SMA specific clinical outcome assessment tool. Singh RN, editor. PLoS One [online serial]. Public Library of Science; 2017;12:e0172346. Accessed at: https://dx.plos.org/10.1371/journal.pone.0172346. Accessed September 20, 2020.

34. Cano SJ, Mayhew A, Glanzman AM, et al. Rasch analysis of clinical outcome measures in spinal muscular atrophy. Muscle and Nerve [online serial]. NIH Public Access; 2014;49:422–430. Accessed at: http://www.ncbi.nlm.nih.gov/pubmed/23836324. Accessed August 20, 2019.

35. Montes J, Dunaway Young S, Mazzone ES, et al. Nusinersen improves walking distance and reduces fatigue in later-onset spinal muscular atrophy. Muscle and Nerve. 2019;60:409–414.

36. Elsheikh B, King W, Peng J, et al. Outcome measures in a cohort of ambulatory adults with spinal muscular atrophy. Muscle Nerve [online serial]. John Wiley and Sons Inc.; 2020;61:187–191. Accessed at: https://onlinelibrary.wiley.com/doi/abs/10.1002/mus.26756. Accessed January 9, 2021.

37. Werlauff U, Steffensen BF. The applicability of four clinical methods to evaluate arm and hand function in all stages of spinal muscular atrophy type II. Disabil Rehabil [online serial]. Informa Healthcare; 2014;36:2120–2126. Accessed at: https://pubmed.ncbi.nlm.nih.gov/24579651/. Accessed January 9, 2021.

38. Jochmann E, Steinbach R, Jochmann T, et al. Experiences from treating seven adult 5q spinal muscular atrophy patients with Nusinersen. Ther Adv Neurol Disord [online serial]. SAGE Publications Ltd; 2020;13. Accessed at: https://pubmed.ncbi.nlm.nih.gov/32180828/. Accessed December 25, 2020.

39. Trucco F, Ridout D, Scoto M, et al. Respiratory Trajectories in Type 2 and 3 Spinal Muscular Atrophy in the iSMAC Cohort Study. Neurology [online serial]. Ovid Technologies (Wolters Kluwer Health); 2021;96:e587–e599. Accessed at: https://pubmed.ncbi.nlm.nih.gov/33067401/. Accessed January 5, 2021.

40. Bakker LA, Schröder CD, Tan HHG, et al. Development and assessment of the inter-rater and intra-rater reproducibility of a self-administration version of the ALSFRS-R. J Neurol Neurosurg Psychiatry [online serial]. BMJ Publishing Group; 2020;91:75–81. Accessed at: https://pubmed.ncbi.nlm.nih.gov/31558653/. Accessed April 11, 2021.

41. Elsheikh B, Prior T, Zhang X, et al. An analysis of disease severity based on SMN2 copy number in adults with spinal muscular atrophy. Muscle and Nerve. 2009;40:652–656.

42. Fournier CN, Bedlack R, Quinn C, et al. Development and Validation of the Rasch-Built Overall Amyotrophic Lateral Sclerosis Disability Scale (ROADS). JAMA Neurol. Epub 2019.

43. van Eijk RPA, de Jongh AD, Nikolakopoulos S, et al. An old friend who has overstayed their welcome: the ALSFRS-R total score as primary endpoint for ALS clinical trials. Amyotroph Lateral Scler Front Degener [online serial]. Informa UK Limited; Epub 2021 Feb 2.:1–8. Accessed at: https://www.tandfonline.com/doi/full/10.1080/21678421.2021.1879865. Accessed February 10, 2021.

44. Vázquez-Costa JF, Hervás D. Minimal detectable change and minimal clinically important difference in spinal muscular atrophy patients. Eur J Neurol. Epub 2021.:1–2.

45. Madruga-Garrido M, Vázquez-Costa JF, Medina-Cantillo J, et al. Design of a Non-Interventional Study to Validate a Set of Patient- and Caregiver-Oriented Measurements to Assess Health Outcomes in Spinal Muscular Atrophy (SMA- TOOL Study). Neurol Ther. Epub 2021.

